# Severe COVID-19 patients display a back boost of seasonal coronavirus-specific antibodies

**DOI:** 10.1101/2020.10.10.20210070

**Authors:** Brenda M. Westerhuis, Muriel Aguilar-Bretones, Matthijs P. Raadsen, Erwin de Bruin, Nisreen M.A. Okba, Bart L. Haagmans, Thomas Langerak, Henrik Endeman, Johannes P.C. van den Akker, Diederik A.M.P.J. Gommers, Eric C.M. van Gorp, Barry H.G. Rockx, Marion P.G. Koopmans, Gijsbert P. van Nierop

## Abstract

Severe acquired respiratory syndrome coronavirus-2 (SARS-CoV-2) is the cause of coronavirus disease (COVID-19). In severe COVID-19 cases, higher antibody titers against seasonal coronaviruses have been observed than in mild cases. To investigate antibody cross-reactivity as potential explanation for severe disease, we determined the kinetics, breadth, magnitude and level of cross-reactivity of IgG against SARS-CoV-2 and seasonal CoV nucleocapsid and spike from 17 severe COVID-19 cases at the clonal level. Although patients mounted a mostly type-specific SARS-CoV-2 response, B-cell clones directed against seasonal CoV dominated and strongly increased over time. Seasonal CoV IgG responses that did not neutralize SARS-CoV-2 were boosted well beyond detectable cross-reactivity, particularly for HCoV-OC43 spike. These findings support a back-boost of poorly protective coronavirus-specific antibodies in severe COVID-19 patients that may negatively impact *de novo* SARS-CoV-2 immunity, reminiscent of original antigenic sin.

## Introduction

The introduction of the zoonotic severe acute respiratory syndrome coronavirus 2 (SARS-CoV-2) has led to a pandemic of acute respiratory tract disease termed COVID-19^1^. The majority of COVID-19 patients experience mild symptoms including fever, sore throat, cough, dyspnea, anosmia, ageusia and myalgia, none of which can be considered specific to this infection^2^. In some patients, SARS-CoV-2 infection leads to acute respiratory distress syndrome (ARDS) with prolonged disease course requiring treatment at an intensive care unit (ICU) and resulting in a high mortality rate^2–4^.

Risks for developing ARDS and poor disease outcome are older age and factors consistent with immune activation^5^. Correlates of protection for severe COVID-19 are not defined in humans but antibodies likely are important. However, there is concern that suboptimal antibody responses may also be detrimental for COVID-19 outcome^6^. The possibility of immune-mediated disease has been raised as a major concern for SARS-CoV-2 infection and an important consideration is the infection history and resulting immunological background against seasonal coronaviruses (CoV)^6,7^. A lack of knowledge on specific features of the induced host response to SARS-CoV-2 in ARDS patients hampers the development of targeted treatment to prevent or overcome severe disease^5^. Therefore, there is an urgent need for detailed insight in the SARS-CoV-2 immune response in the context of a CoV-experienced immune system.

For other viruses, several mechanisms have been described on how pre-existing immunity to antigenically related viruses affect the outcome of infection. First, for influenza and dengue viruses the immunological imprint of the initial infection dominates the host response towards subsequent related infections for life^8,9^. This mechanism, termed “original antigenic sin” (OAS), relates to the propensity of adaptive immunity to preferentially fight pathogens based on memory recall. Consequently, clones primed to target a specific epitope may dominate the response to target a new slightly different cross-reactive epitope and have reduced affinity and functionality, e.g. a poor neutralization potential for the new infection^8–10^. Moreover, OAS has also been linked to a delayed or weakened initiation of a new type-specific response^10^.

Second, for influenza virus increased antibody titers targeting viruses far beyond the response against the first encountered antigen alone were observed^11,12^. The observed boost in neutralization titers was stratified on antigenic seniority, with highest titer increase towards more senior antigens^12^. Also, a broad “back boost”, extending beyond cross-reactivity, after infection or vaccination has been shown^11,13^. This phenomenon has not yet been described for any other viral infection but may have strong implications for vaccine design and the response to infection. Although a back-boost by vaccination may be useful to maintain broad immunity to preceding strains, it may negatively affect the development of a newly induced immune response, similar to OAS^9–11^.

Structural homology between the ectodomain of SARS-CoV-2 Spike (S, SARS2-S_ECTO_) or nucleocapsid protein (N, SARS2-N) with those of other endemic human seasonal CoV such as hCoV-229E, -NL63, -HKU1 and -OC43 or recent epidemic strains such as SARS-CoV and Middle-East respiratory syndrome coronavirus (MERS-CoV) suggests that memory B-cells capable of expressing cross-reactive antibodies may pre-exist in patients^14,15^. This is exemplified by the cross-reactive antibody response towards SARS-CoV-2 in SARS-CoV infected individuals due to high sequence homology (88.6% shared amino acids in N and 69.2% in S)^14,16^. Structural homology between SARS-CoV-2 and seasonal CoV is lower^17^, yet these viruses are highly prevalent^16^. Repeated exposure to seasonal CoV may therefore also substantially contribute to developing a SARS-CoV-2 reponse^18^.

A strong SARS2-N and SARS2-S IgM and IgG response is mounted in the vast majority of infected individuals targeting the receptor binding domain (S_RBD_), the outer-(S_1_) and inner domains (S_2_) to various extent^19^. In severe COVID-19 cases this response is dominated by IgG targeting the S_2_ domain^16,19^. There was no evidence of substantial IgG cross-reactivity towards SARS-CoV-2 in healthy controls^16,19^. In contrast, COVID-19 patients displayed elevated serum IgG titers against the majority of seasonal CoV N and S antigens which suggests that the IgG responses against these viruses are correlated by undefined mechanisms. Strikingly, high serum OC43-S_ECTO_ IgG titers are associated with COVID-19 disease severity, which might be related to stretches of amino acid sequence homology in S_2_ ^16,17^. Together, these studies suggest that high antibody titers to seasonal CoV may associate with poor disease outcome in COVID-19 patients and support the hypothesis that the immunological background is key. A limitation of serological studies is that monoclonal IgG cross-reactivity patterns cannot be defined and the functional contribution of seasonal CoV-specific IgG clones to immune protection or immunopathogenesis remains largely unknown.

At the clonal level, it was shown that pre-existing SARS-CoV-2 cross-reactive IgG memory B-cells predominantly target the SARS-CoV-2 S_2_ with limited virus neutralization (VN) potential^20^. Besides a shared epitope that has been described in S_2_ of SARS-CoV-2 and OC43^21^, little is known on seasonal CoV IgG cross-reactivity patterns with SARS-CoV-2 and their functional contribution. Pre-existing CoV-specific B-cells may provide protection from SARS-CoV-2 infection by rapid production of cross-reactive antibodies that aid in viral clearance, e.g. by a faster rise of cross-neutralizing antibodies from memory recall compared to initiating a primary humoral response^22,23^. However, based on observations in other viral infections, pre-existing immunity may also promote pathology^7^.

Here, we longitudinally enumerated and categorized type-specific and cross-reactive circulating B-cell clones targeting an array of N and S antigens from a broad selection of human CoV in order to gain insight in immune kinetics, magnitude, breadth and function towards SARS-CoV-2 in COVID-19 patients admitted to the intensive care unit (ICU).

## Methods

### Patient characteristics and clinical specimen

PCR-confirmed COVID-19 patients^24^ admitted to the ICU at the Erasmus Medical Center with acute respiratory distress syndrome (ARDS) were included in a biorepository study (MEC-2017-417) and samples were analyzed according to the SARS-CoV-2 protocol (MEC-2020-0222) with approval from the medical ethical committee of the Erasmus MC. Personal or deferred informed written consent was obtained after recovery or from a legal representative, partner or family member of the participant respectively, all in compliance with the Declaration of Helsinki. Plasma and peripheral blood mononuclear cells (PBMC) were isolated from EDTA vacutainer blood collection tubes (BD biosciences) aliquoted and cryopreserved as described elsewhere^25^. Weekly serum and EDTA blood samples for PBMC isolation were collected where the initial sample was collected within 2 days after admittance to the ICU until patients were released from the ICU. From patients 1–6, samples from the first three weeks were included in this study for longitudinal analysis. Additional serum samples from later time points were included in the study if available. Additionally, 11 cross-sectionally sampled COVID-19 patients were selected based on differential clinical outcome of infection. Six succumbed to, and five recovered from SARS-CoV-2 infection (Table 1).

**Table 1.** Patient characteristics.

| Patient ID | Gender | Age group | Days post onset symptoms at inclusion | Duration stay ICU (days) | Survival<sup>1</sup> | Thrombotic complication | Comorbidities |
| --- | --- | --- | --- | --- | --- | --- | --- |
| 1 | M | 70-80 | 10 | 50 | Y | Y | None |
| 2 | F | 50-60 | 4 | 26 | Y | Y | Hypertension |
| 3 | M | 50-60 | 5 | 17 | Y | N | Lung disease, Malignancy |
| 4 | F | 60-70 | 9 | 21 | Y | N | None |
| 5 | M | 60-70 | 5 | 31 | Y | N | Cardiac disease, Hypertension |
| 6 | M | 50-60 | 12 | 45 | Y | Y | Cardiac disease, Hypertension, Vascular disease, Type 2 diabetes, Lung disease |
| 7 | M | 70-80 | 15 | 15 | Y | N | Vascular disease, Type 2 diabetes |
| 8 | M | 60-70 | 14 | 8 | Y | N | Malignancy |
| 9 | M | 50-60 | 15 | 10 | Y | Y | Type 2 diabetes |
| 10 | F | 70-80 | 20 | 13 | Y | N | Hypertension |
| 11 | F | 70-80 | 18 | 10 | N | Y | Type 2 diabetes |
| 12 | M | 20-30 | 9 | 10 | N | N | None |
| 13 | F | 70-80 | 16 | 8 | N | Y | Cardiac disease, Vascular disease |
| 14 | M | 60-70 | 33 | 26 | N | Y | Hypertension |
| 15 | M | 50-60 | 8 | 4 | N | Y | Hypertension |
| 16 | M | 70-80 | 29 | 11 | Y | Y | None |
| 17 | M | 60-70 | 13 | 5 | Y | N | Cardiac disease, Type 2 diabetes |

<sup>1</sup>Patients were followed up for at least 60 days.

### B-cell profiling

B-cells were isolated from cryopreserved PBMC using the EasySep human CD19 positive selection kit (Stem Cell technologies) according to manufacturer’s instructions. Oligoclonal cultures of 100-300 CD19+ B-cells per well were seeded in 96 wells U-bottom plates in AIM-V AlbuMAX medium supplemented with 10% fetal bovine serum, penicillin-streptomycin (all Invitrogen) and β-mercaptoethanol (Sigma) (B-cell medium). Oligoclonal B-cell cultures were stimulated using 1000 L-CD40L cells that were growth arrested by 40Gray γ-irradiation, 50 U/ml IL-2 (Novartis), 10 ng/ml IL-10 (Peprotech), 25 ng/ml IL-21 (Peprotech) and 1 µg/ml R848 (Invivogen) for 48 hours and subsequently cultured for 12 days in B-cell medium supplemented with 25ng/ml IL-21. Culture supernatants were harvested and reactivity of secreted IgG was determined using protein microarray analysis.

### Protein micro-array analysis

Reactivity of secreted IgG in oligoclonal culture supernatants towards an array of CoV N and S proteins was analyzed using protein microarray analysis (PMA) as described elsewhere^26^. Paired sera of the respective PBMC samples were included in the analysis. Microarray slides were scanned using a Powerscanner (Tecan, Switzerland). Background was determined for each spot and mean fluorescence intensity signal (MFI, range 1 – 65,000) of two spots was calculated for each serum or culture supernatant. For B-cell culture supernatants a cut-off was set at 1,000 MFI for all antigens based on the highest signal of non-reactive cultures. Serum IgG titers were calculated using 4-parameter logistic regression with inflection point as titer^27^.

### Virus neutralization assays

Virus neutralization was determined by plaque reduction neutralization titers as previously described^17^.

### Statistical analysis

Serum IgG and virus neutralization titers and mean fluorescent intensity values of protein microarray analysis were ^2^Log transformed for regression analysis. P-values below 0.05 were considered statistically significant. All analyses were performed using ‘R’ and Graphpad Prism 7 as previously described^18,27^.

## Results

### Clinical characteristics of study participants

Seventeen severe COVID-19 patients with moderate to severe ARDS were included in this study, shortly after being admitted to the ICU or the Erasmus MC, Rotterdam (Table 1). From six patients we included three or more longitudinal samples. All patients were tested RT-PCR positive for SARS-CoV-2 and required mechanical ventilation during their stay at ICU. Five females (mean age = 67.2y, range = 54-75y), and 12 males were included in the study (mean age = 59.9y, range = 29-77y). Five patients died, nine recovered and three were transferred to different hospitals for further treatment. Thirteen out of seventeen patients suffered from one or more comorbidities, of which hypertension (n=6), and type 2 diabetes mellitus (n=5), were the most common. Nine patients experienced thrombotic complication during their stay at the ICU (Table 1).

### Serum IgG titers towards a broad range of coronavirus N and S antigens increased after SARS-CoV-2 infection in severe COVID-19 patients

All patients mounted a strong SARS-CoV-2 immune response shown by rising IgG titers against SARS2-S_ECTO_, SARS2-S_1_ and SARS2-S_RBD_ which peaked around 4 weeks post onset of clinical symptoms (Figure 1A and Supplementary figure 1A for individual patients). These patients were assumed never exposed to SARS-CoV and MERS-CoV. Nevertheless, there was a strong IgG response towards all SARS-CoV antigens. Given the high structural similarities between N and S of SARS-CoV and SARS-CoV-2^16,28^, likely a cross-reactive response was mounted. The increase in MERS-S_ECTO_ IgG, but not MERS-N and MERS-S_1_ IgG and highest amino acid homology within S_2_ with SARS-CoV-2 suggested this cross-reactive response targeted MERS-S_2_ ^17^.

**Figure 1.**
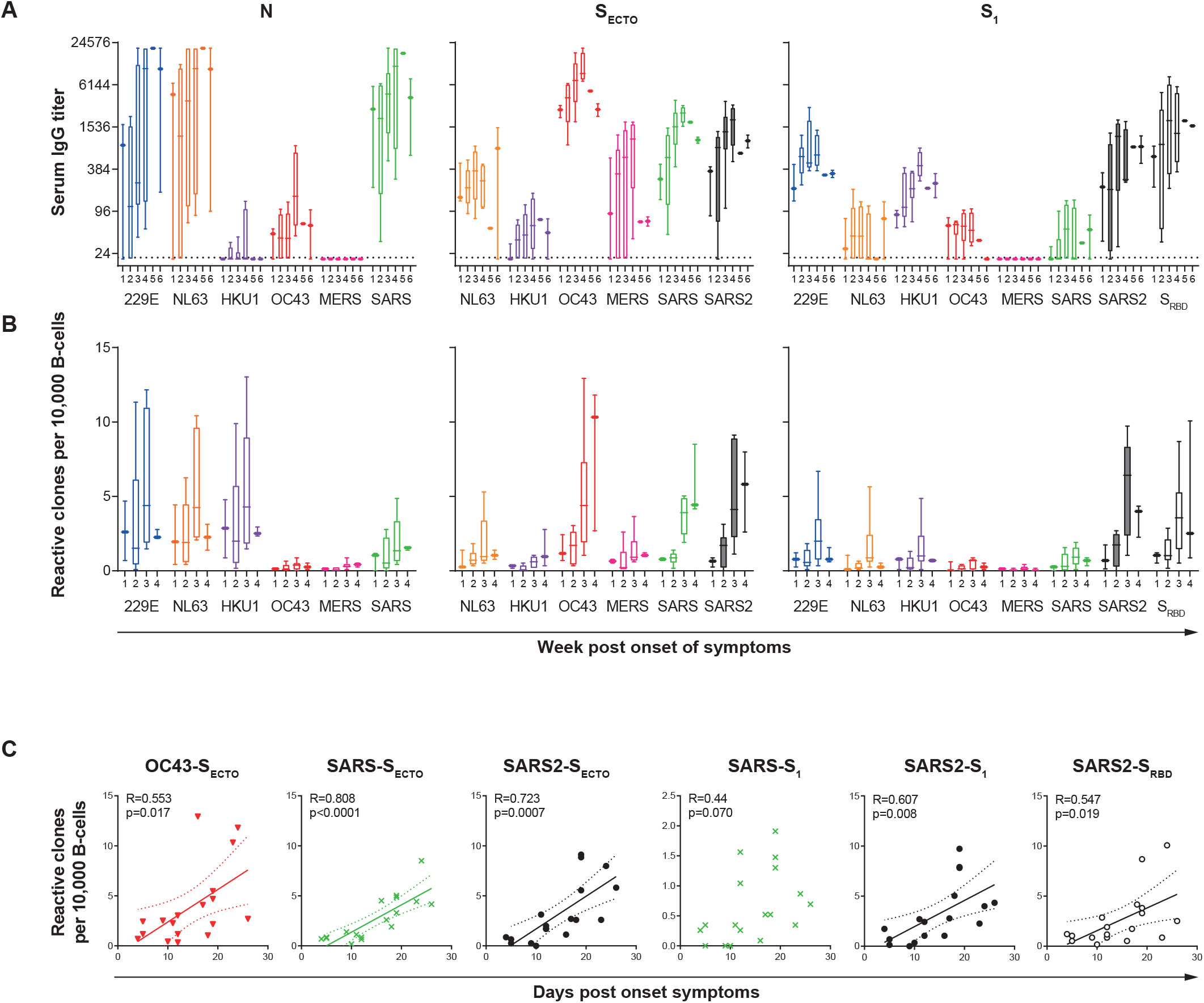
Severe COVID-19 patients generate a strong SARS-CoV-2-specific IgG response and display an increasing IgG response towards other coronaviruses. **A**) Longitudinal serum IgG responses of six severe COVID-19 patients sampled three times with weekly intervals, grouped per week number after onset of symptoms, towards a panel of nucleocapsid proteins (N), the ecto-(S_ECTO_) and head-domains (S_1_) of spike or the SARS-CoV-2 (SARS2) receptor binding domains (S_RBD_). A SARS-CoV-2 response is mounted in all patients, together with a boost of seasonal human 229E, NL63, HKU1, OC43 and emerging MERS, SARS and SARS2 coronavirus antigens. Dotted line shows the assay background. **B**) Enumeration of paired *in vitro* stimulated peripheral blood-derived B-cells at limiting density with IgG reactivity towards coronavirus antigens, normalized for the number of analyzed B-cells. Box represents median, upper and lower quartile. Whiskers show range. **C**) OC43-S_ECTO_, SARS-S_ECTO_ SARS2-S_ECTO_/S_1_ and spike receptor binding domain (S_RBD_) specific B-cell clones show a synergistic linear outgrowth in the first four weeks after SARS-CoV-2 infection. Pearson correlation test, solid line shows regression, dotted line shows the 95% confidence interval of the best-fit line.

Besides the SARS-CoV-2 IgG response, serum IgG reactivity towards various, but not all seasonal CoV antigens, increased in longitudinally analyzed COVID-19 patients (Figure 1A and Supplementary figure 1A). All patients showed IgG reactivity towards at least one of the 229E, NL63, HKU1 or OC43 antigens (Supplementary figure 1A). Potential IgG cross-reactivity in serum responses refutes any firm conclusions on previous exposure against these CoV. For S, OC43-S_ECTO_ IgG titers were immunodominant at inclusion in all six patients and titers increased in five out of six patients during follow-up.

### Severe COVID-19 patients showed outgrowth of circulating B-cells targeting a broad range of coronaviruses

To analyze the kinetics of the IgG response we longitudinally enumerated circulating B-cell clones targeting CoV proteins using B-cell profiling on six severe COVID-19 patients (Supplementary figure 1B). Paired CD19+ B-cells isolated from PBMC of COVID-19 patients were stimulated oligoclonally *in vitro* at a limiting density that was empirically tested to give a clonal B-cell response. Supernatants were individually screened for IgG reactivity towards all available CoV antigens using PMA. The number of reactive B-cells was normalized for the number of screened B-cells in order to compare frequencies between different samples (Figure 1B). Overall, the frequency and total reactivity of *in vitro* stimulated peripheral blood-derived B-cells was representative for the total serum IgG reactivity (Figure 1B and supplementary figure 1B). Similar to serum IgG reactivity, all patients showed a strong increase in SARS-CoV-2 reactive B-cells and the number of B-cell clones reactive towards various seasonal CoV antigens increased after SARS-CoV-2 infection. However, the frequency of HKU1-N reactive B-cells was higher than expected based on the low serum reactivity. Potentially, this difference reflects a different level of B-cell activation *in vitro* and *in vivo*. The linear outgrowth of B-cells reactive towards OC43-S_ECTO_, SARS-S_ECTO_, SARS2-S_ECTO_, SARS2-S_1_ and SARS2-S_RBD_ displayed strikingly similar kinetics in their response (Figure 1C).

### Coronavirus type-specific and cross-reactive IgG clones increased during SARS-CoV-2 infection

The outgrowth of B-cells that do not target SARS-CoV-2 antigens is potentially related to cross-reactivity. Therefore, we analyzed the multiplex PMA profiles of *in vitro* stimulated B-cell cultures to address the level of cross-reactivity of monoclonal IgG. Profiles of CoV antigen-reactive cultures are shown for a representative patient (patient 4, Figure 2). N-reactive clones from this donor showed substantial cross-reactivity between 229E-N, NL63-N and HKU1-N at all time points (Figure 2A). Although individual clones with aberrant cross-reactivity can be identified in these patients, we hypothesize these contribute little to the overall IgG response. A Pearson correlation test was used to calculate the strength and robustness of cross-reactivity between each pair of antigens to identify cross-reactivity patterns that significantly contribute to the induced immune response (Figure 2B). The level of cross-reactivity as calculated by Pearson correlation test varied slightly over time due to the limited number of clones per sample. Overall, the cross-reactivity patterns between 229E-N, NL63-N and HKU1-N became more apparent and statistically more significant when all clones from three time points (n=125) are combined for analysis (Figure 2B). OC43-N and MERS-N clones displayed no cross-reactivity and cross-reactivity between SARS-N and seasonal CoV was considered non-substantial (Figure 2A/B).

**Figure 2.**
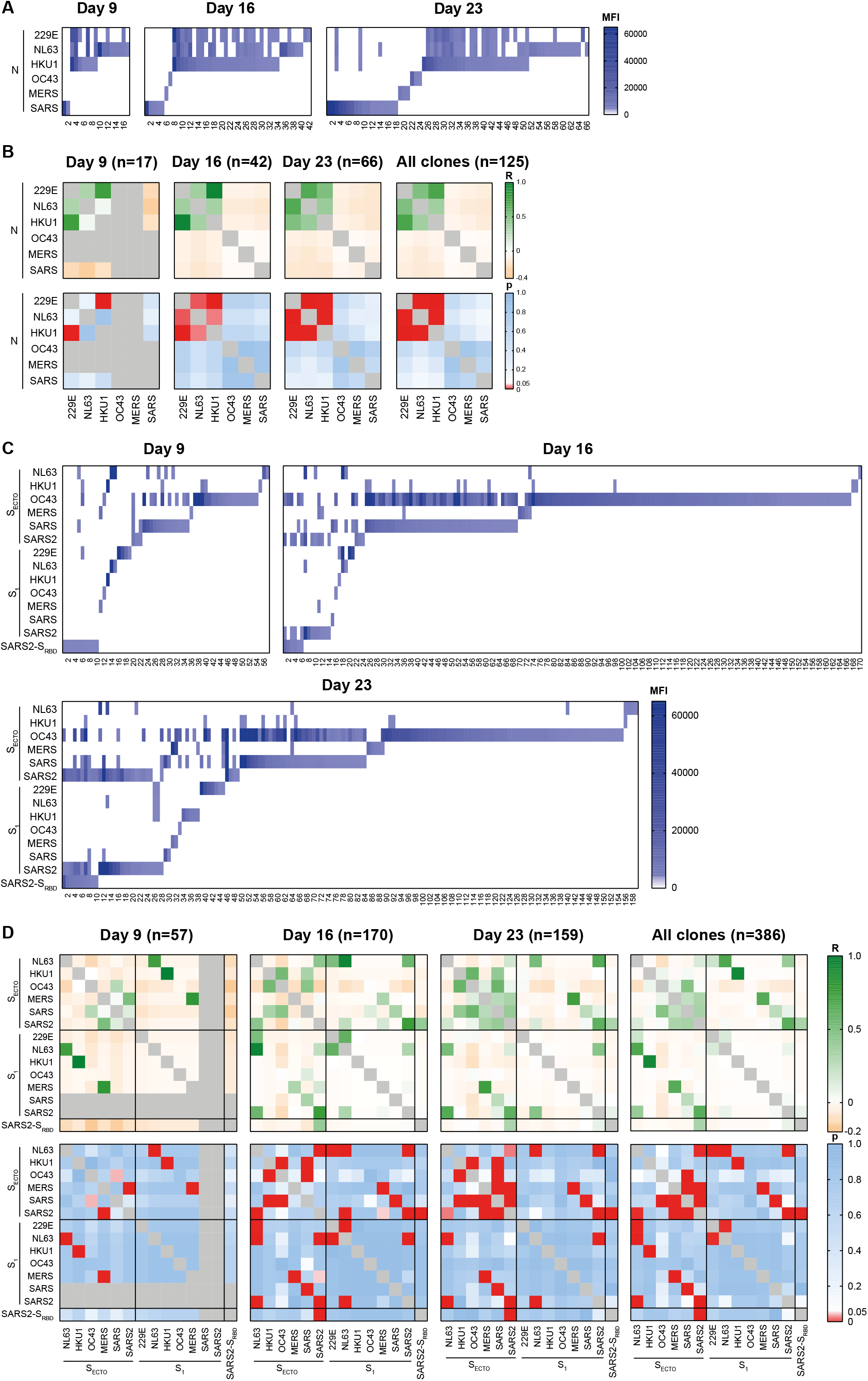
Supernatants of *in vitro* stimulated B-cells of a representative donor shows clonal IgG cross-reactivity patterns towards different coronavirus strains. B-cells were isolated from peripheral blood samples of patient 4 and stimulated *in vitro* in oligoclonal cultures at limiting dilution to analyze IgG reactivity at the clonal level. This representative patient was analyzed at three consecutive weekly intervals starting at 9 days post onset of symptoms. **A**) Heatmaps of the mean fluorescent intensity (MFI) of clonal IgG reactivity towards N of a panel of coronaviruses. The number of single- and cross-reactive N-specific B-cells increased over time after SARS-CoV-2 infection. **B**) Heatmaps of the Pearson regression coefficient (R, top panels) and significance (bottom panels) between N-reactivity of specific B-cell clones shows the level of cross-reactivity at weekly intervals and the clones of all time points combined. **C**) Heatmaps and **D**) regression analysis of S_ECTO_, S_1_ and S_RBD_ reactive IgG clones from the same patient is shown. Positive correlations (green shades) show clones likely cross-react, negative correlations (orange shades) show clones are likely to not cross-react. Significant associations are shown in red shades (p<0.05).

S-reactive IgG clones showed an increasing mostly SARS-CoV-2 type-specific response with increasing numbers of detected SARS2-S_ECTO_ and SARS2-S_1_ B-cell clones. Strong SARS2-S_ECTO_ and SARS2-S_1_ cross-reactivity indicated that mostly shared epitopes within S_1_ were targeted in this patient (Figure 2C and D). The frequency and number of detected SARS2-S_RBD_ clones remained more or less stable, yet cross-reactivity towards SARS2-S_ECTO_ and SARS2-S_1_ increased over time suggesting maturation of the SARS-CoV-2 specific response. The initial lack of cross-reactivity of SARS2-S_RBD_ towards SARS2-S_ECTO_ and SARS2-S_1_ suggested that the epitope(s) targeted by the SARS2-S_RBD_ clones at day 9 were sterically hindered or displayed in an alternative conformation in the larger SARS-CoV-2 S proteins on PMA (Figure 2C). The increasing cross-reactivity suggests clones that target epitopes displayed on the outer surface of S are naturally selected.

Strikingly, there was a dominant OC43-S_ECTO_-specific response that increased over time. This response significantly cross-reacted with HKU1-S_ECTO_ and SARS-S_ECTO_, but not SARS2-S_ECTO_ (Figure 2D). This suggests the outgrowth of OC43-S_ECTO_ clones is not driven by SARS-CoV-2 cross-reactivity. The limited cross-reactivity between OC43-S_ECTO_ and OC43-S_1_ suggested these clones predominantly target S_2_. The correlated response between the paired S_1_ and S_ECTO_ antigens of NL63-, HKU1-, MERS-, SARS- and SARS-CoV-2 validated availability of shared epitopes in both antigens.

The total number of detected S_ECTO_-reactive clones (n=363) outnumbered S_1_-reactive clones (n=85) and showed greater cross-reactivity between different CoV compared to the S_1_. These data suggested clones targeting S_2_ are immunodominant and more cross-reactive in this donor.

For the other five longitudinally analyzed donors, similar patterns were observed with *i)* high levels of cross-reactivity between 229E-N, NL63-N and HKU1-N but not SARS-N and MERS-N *ii)* a mostly SARS-CoV-2 type-specific response that strongly increased over time, *iii)* an OC43-S_2_-specific response that increased over time with limited cross-reactivity towards SARS-CoV-2. Notably, the N-reactivity of 1/6 patients showed increased cross-reactivity between OC43-N and other seasonal strains (Supplementary figure 6). OC43-S_ECTO_-specific B-cells were immunodominant in 3/6 patients (Supplementary figure 1B) and the S_2_ response was immunodominant in 2/6 patients (Supplementary figures 2–6).

### N-IgG selectively cross-reacted between seasonal coronaviruses while S-IgG showed complex cross-reactivity patterns involving all strains

In order to determine the overall pattern of IgG cross-reactivity between N and S of different CoV in severe COVID-19 patients we pooled all available N (n=2039, Figure 3A) and S-reactive IgG clones (n=2699, Figure 3B) from all 17 included donors. For N, Pearson correlation analysis validated that 229E-N, NL63-N and HKU1-N IgG clones most substantially cross-reacted (range R=0.52 – 0.72, all p<0.0001, Figure 3C). Additionally, weak yet highly significant correlations between OC43-N and other seasonal CoV strains were observed (range R=0.066 – 0.14, all p<0.003). Conversely, no significant cross-reactivity was detected between seasonal and emerging CoV indicating different epitopes to be involved for these recent zoonotic strains (Figure 3C).

**Figure 3.**
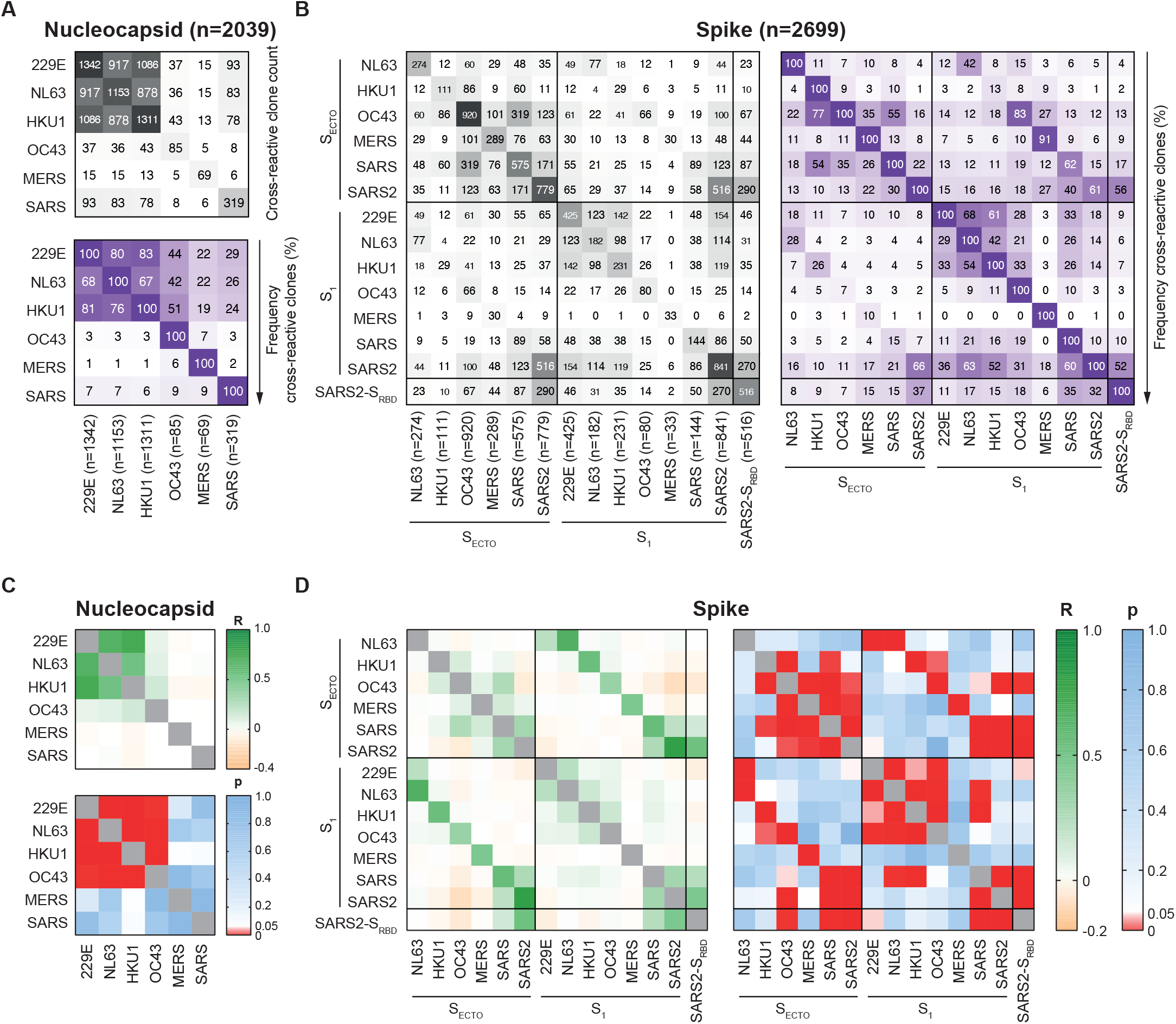
Combined analysis of reactive clones from all donors shows cross-reactivity patterns between human seasonal and emerging coronaviruses. Reactivity of N (n=2039) and S-reactive IgG clones (n=2699) identified in 17 severe COVID-19 patients was analyzed. **A**) Heatmaps show the relative amount (top panel, grey shades) and number of IgG clones reactive towards at least one (top left to bottom right diagonal) or two N antigens and the relative frequency of cross-reactivity towards these antigens (bottom panel, purple shades, numerator is shown in clone count heatmap, denominator is indicated on x-axis). **B**) Relative amount and number (left panel) and frequency (right panel) of S-cross-reactive IgG clones. **C**) Pearson regression analysis of 2039 N reactive clones shows significant cross-reactivity between all human seasonal coronaviruses. **D**) Regression analysis of 2699 S_ECTO_, S_1_ and S_RBD_ reactive IgG clones shows broad cross-reactivity between human seasonal and emerging coronavirus strains.

The combined analysis of all S-IgG clones showed complex cross-reactivity patterns, including endemic seasonal and epidemic zoonotic strains (Figure 3D). All SARS-CoV and SARS-CoV-2 S antigens positively correlated (range R=0.14 – 0.80, all p<0.0001). Additionally, all corresponding S_1_ and S_ECTO_ antigens positively correlated (range R=0.35 – 0.80, all p<0.0001) validating availability of shared epitopes in paired antigens for all CoV strains.

Our cohort showed there was broad cross-reactivity within S_1_ between 229E-, NL63-, HKU1- and OC43-CoV but not MERS-CoV. For MERS-CoV significant cross-reactivity was restricted to S_ECTO_, suggesting cross-reactive epitopes are mostly located in S_2_ (Figure 3D).

### Correlated clonal IgG reactivity patterns showed in vivo expansion of OC43-S_ECTO_ specific B-cells beyond detectable cross-reactivity

Given the synergistic outgrowth of OC43-S_ECTO_ and SARS-CoV and SARS-CoV-2 S-reactive B-cells in severe COVID-19 patients (Figure 1C) one would presume that SARS-CoV-2 cross-reactivity drives *in vivo* expansion of OC43-S_ECTO_-specific B-cells. However, cross-reactivity between OC43-S_ECTO_ and SARS-CoV-2 S antigens showed significant weak negative correlations (range R=-0.038 – −0.088, range p=4,7×10^−6^ – 0.014) suggesting that OC43-S_ECTO_ specific clones are unlikely to cross-react with SARS-CoV-2 S in severe COVID-19 patients (Figure 3B). Indeed, the majority of OC43-S_ECTO_ clones (752 out of 920 clones, 82%) did not cross-react with SARS-CoV-2 indicating these were initiated prior to SARS-CoV-2 infection. Due to lack of a pre-sample we cannot draw firm conclusions on pre-existence of SARS-CoV-2 cross-reactive clones in this cohort.

Longitudinal analysis of OC43-S_ECTO_ clones in all 17 patients, grouped per week number post onset of symptom showed the average relative number of clones that cross-reacted with SARS-CoV-2 S antigens showed a moderate increase over 4 weeks (13 – 21%). The OC43-S_ECTO_-reactive clones that cross-reacted increasingly recognized epitopes in SARS2-S_1_ and SARS2-S_RBD_ indicating there was natural selection of SARS-CoV-2 cross-reactive clones specific for the outer S domains (Figure 4).

**Figure 4.**
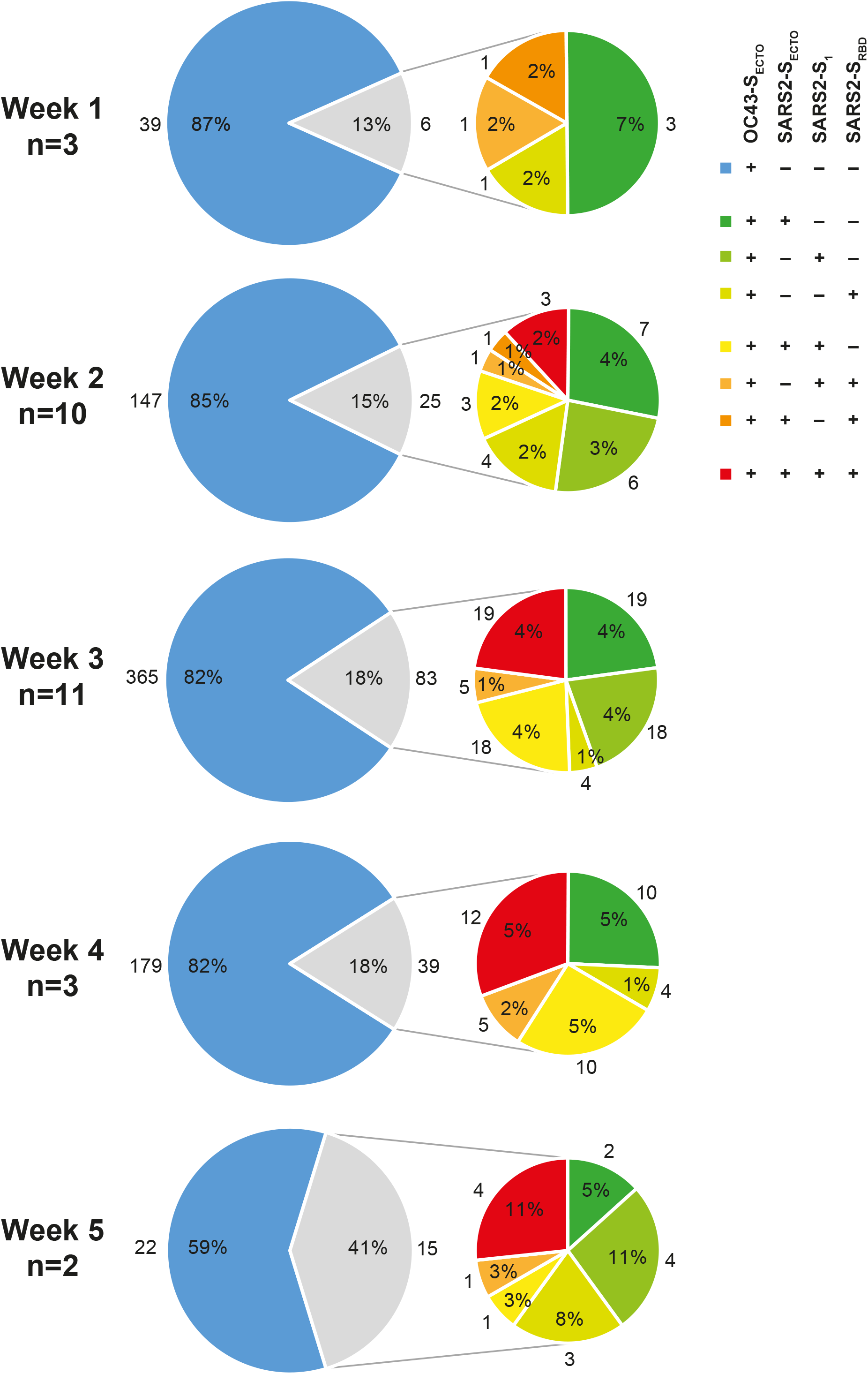
OC43-S_ECTO_-reactive clones show minor yet antigenically evolving cross-reactivity towards SARS-CoV-2. All OC43-S_ECTO_-reactive clones (n=920) from 17 patients were stratified by week number post onset of clinical symptoms. The proportion of single OC43-S_ECTO_ (blue) and SARS-CoV-2 cross-reactive clones (grey) is shown (left pie). The cross-reactivity towards a single (green colors), dual (yellow and orange colors) and three SARS-CoV-2 antigens (red color) is depicted (right pie). The number of patients analyzed is indicated on the left. The frequency and number of contributing clones for each cluster is shown inside and adjacent to each part, respectively.

### No evidence for functional contribution of OC43-S_ECTO_-specific IgG in virus neutralization

To determine what CoV-specific IgG functionally contribute to clearing SARS-CoV-2 infection via virus neutralization (VN) we correlated serum S reactive IgG to serum SARS-CoV-2 VN titers. All SARS-CoV and SARS-CoV-2 S antigens selectively positively correlated with serum SARS-CoV-2 VN titers but none of the other CoV S antigens. This is congruent with SARS-CoV/SARS-CoV-2 cross-reactivity patterns and other studies showing that S is the dominant antigenic target of neutralizing clones^29,30^. Notably, no correlation was detected between VN and serum OC43-S_ECTO_ and OC43-S_1_ IgG titers (Figure 5A).

**Figure 5.**
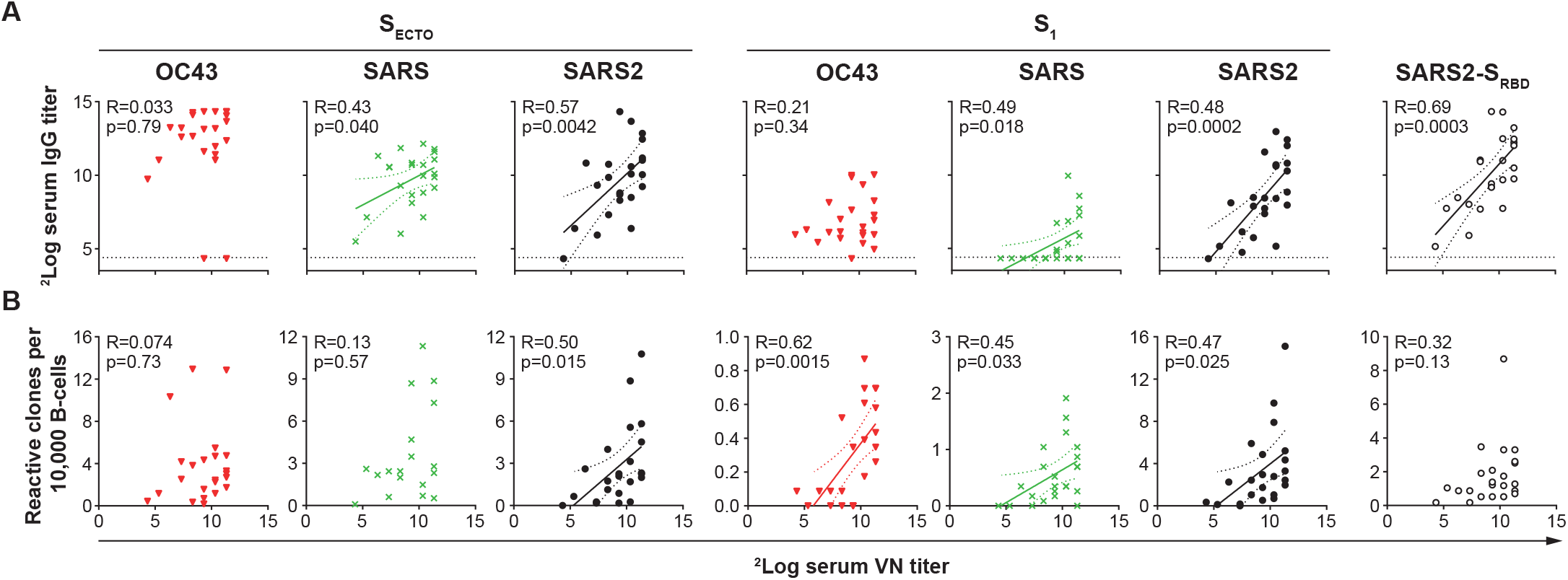
Serum IgG titers or normalized B-cell counts reactive towards OC43-S_1_, SARS-S_ECTO_/S_1_ and SARS2-S_ECTO_/S_1_ correlate with serum SARS-CoV-2 virus neutralization titers, but not OC43-S_ECTO_. **A**) Linear correlation between ^2^Log transformed serum IgG and SARS-CoV-2 virus neutralization (VN) titers shows that only SARS-CoV and SARS-CoV-2 spike-reactive IgG associate with VN. Horizonal dotted line shows the background of the assay. **B**) Normalized counts of B-cells specific for SARS-S_ECTO_, OC43-S_1_, SARS-S_1_ and SARS2-S_1_ selectively correlate with VN. 22 Serum samples and corresponding B-cell cultures of 17 patients were included in these analyses. Solid line depicts the best-fit regression coefficient. Dashed line shows the 95% confidence of the best-fit line.

To determine what circulating S-specific B-cells contribute to VN, the normalized S-reactive B-cell counts were correlated with paired serum SARS-CoV-2 VN titers. Similar to serum IgG titers, SARS2-S_ECTO_, SARS-S_1_ and SARS2-S_1_ correlated with serum VN titers. Contrasting to serum titers, OC43-S_1_ B-cell numbers showed a positive correlation with VN. Notably, the relatively low frequency of OC43-S_1_-reactive B-cells (<0.01% of screened B-cells) suggested the potential functional contribution of these clones to VN *in vivo* was limited. Contrasting to serum IgG, SARS2-S_RBD_ B-cell counts did not reach significance in correlation with VN titers (p=0.13), which may be related to the limited number of samples analyzed (n=22). Notably, neither OC43-S_ECTO_-specific serum IgG nor circulating specific B-cells correlated with serum SARS-CoV-2 VN, which suggests that the strong OC43-S_ECTO_ response had no substantial contribution to SARS-CoV-2 neutralization (Figure 5B).

## Discussion

Multiplex IgG analysis of culture supernatants of *in vitro* activated B-cell cultures at limiting dilutions allowed us to *i)* categorize strain-specific from cross-reactive monoclonal IgG, ii) distinguish pre-existing clones targeting seasonal CoV from newly induced SARS-CoV-2-type specific and cross-reactive clones, *iii)* perform in-depth analysis of humoral immune kinetics in severe COVID-19 patients. Together with paired serological analysis these data gave detailed insight in the magnitude, breadth and cross-reactivity patterns of the SARS-CoV-2 induced immune response in severe COVID-19 patients.

Severe COVID-19 patients displayed an evolving SARS-CoV-2 specific IgG response that correlated with the increasing SARS-CoV-2 neutralization potential of serum during disease progression. Additionally, these patients displayed a strong rise of pre-existing seasonal CoV-specific clones that did not correlate with SARS-CoV-2 neutralizing titers, which is suggestive of OAS. Based on this mechanism, one would expect that cross-reactivity to SARS-CoV-2 would drive outgrowth of pre-existing seasonal CoV-specific B-cells. However, here we observed a strong outgrowth of clones that selectively targeting seasonal CoV. Using Pearson regression analysis, we were able to distinguish rare from substantial cross-reactivity patterns. Although clones were identified that showed SARS-CoV-2 cross-reactivity, these had a minor contribution to the overall SARS-CoV-2 response.

For N, very limited cross-reactivity was observed between seasonal and emerging CoV strains. Even though we could not confirm this for SARS-CoV-2, the high level of SARS2-N and SARS-N sequence homology (88.6%) and reported strong correlation between serum SARS2-N and SARS-N IgG titers^16^ suggests SARS-N may be used as a proxy of SARS2-N. This suggests the outgrowth of 229E-N, NL63-N and HKU1-N reactive IgG clones was not selectively driven by cross-reactivity.

The outgrowth of seasonal CoV-reactive B-cell clones was most striking for OC43-S_ECTO_. OC-43-S_ECTO_-reactive clones outnumbered others in the majority of patients and showed a synergistic outgrowth with the SARS-CoV-2-reactive B-cells. Of OC43-S_ECTO_ reactive clones only a minor fraction cross-reacted with OC43-S_1_ (66 out of 920 clones, 7%). This suggests the majority of OC43-S_ECTO_ clones target S_2_, which corresponds with previous studies^16,21^. The fraction of OC43-S_ECTO_ clones that cross-reacted with SARS-CoV-2 was limited (168 out of 920 clones, 18%) but for the clones that did, reactivity was increasingly directed towards SARS2-S_1_ and SARS2-S_RBD_ and not S_2_. As IgG targeting S_1_ and S_RBD_ confer the strongest VN potential, these OC43-S_ECTO_ clones contribute to evolving SARS-CoV-2-specific IgG response^23,31^.

OC43-S_2_ clones potentially have low-affinity reactivity towards SARS-CoV-2, that is below the detection limit of protein microarray, which drives expansion of OC43-S_ECTO_ clones. This is of interest because low affinity antibodies, with consequentially a poor neutralizing potential, can mediate engagement of Fc receptors (FcRs) on innate immune cells and promote viral entry through a mechanism termed antibody-dependent enhancement (ADE). This may either result in productive infection and increased viral spread – this was shown for flaviviruses^32^ – or promote inflammation and tissue damage, as was described for SARS-CoV^33–35^. However, the level of OC43-S_ECTO_ clones that cross-react with SARS2-S_2_ remained stable over time, as determined by the OC43-S_ECTO_ clones that selectively cross-react with SARS2-S_ECTO_ (Week 1, 7%; week 2, 4 %; week 3, 4% and week >4, 5%). If OC43-S_ECTO_ clones were selectively expanding due to undetected low affinity cross-reactivity, one would expect that clones that have detectable cross-reactivity to have a growth advantage. The reason for this is that the clonal selection of B-cells is driven by the affinity of the B-cell antigen receptor and SARS-S_ECTO_-specific T_H_-cell-mediated linked recognition of antigens, the presence of which has been confirmed in an largely overlapping patient cohort^25^.

We find no evidence of a functional contribution of OC43-S_ECTO_ reactive IgG to SARS-CoV-2 VN. OC43-S_1_ specific clones did correlate with SARS-CoV-2 VN titers although these clones represent only a minor fraction of OC43-CoV specific clones. While it is possible that OC43-S_ECTO_ specific IgG restrict viral replication by indirect mechanisms, they may also play no role, or even have a detrimental effect by delaying the development of a type-specific response or by enhancing immune pathology^36^. The previously reported selective association of increased OC43-S_ECTO_ IgG reactivity in severe versus mild COVID-19 patients supports a potential role in immunopathology^16^. Unfortunately, the limited volume of B-cell culture supernatants was insufficient to perform in-depth analysis of their affinity or function at the clonal level and deserves follow-up studies.

Alternatively, the outgrowth of clones with reactivity towards the highly seroprevalent seasonal strains may relate to a general immune activation induced by a hyper-inflammatory state of the immune system^37^. However, for individual patients, not all CoV-reactive B-cells show similar outgrowth after SARS-CoV-2 infection. Furthermore, the low serum IgG reactivity towards HKU1-N and relatively high frequency of HKU1-N-reactive clones suggests these B-cell are not activated *in vivo*. Together, these data do not support this hypothesis.

Overall, our data suggest that cross-reactivity towards SARS-CoV-2 has a minor contribution to the outgrowth of clones targeting seasonal CoV and *vice versa*. The boosted response involves a broad range of seasonal CoV strains. For influenza virus, the gradual antigenic evolution allows preceding exposures to different virus strains to be chronologically ordered to determine antigen seniority^12^. For seasonal CoV, the order and time window at which individuals were infected or reinfected cannot be deferred. Potentially, the strong and dominant OC43-S_ECTO_ response observed in severe COVID-19 patients relates to antigenic seniority, but this cannot be confirmed. Regardless of the unknown timeline of preceding infections, the outgrowth of seasonal CoV-specific clones is similar to the back-boost that was initially described for influenza^11^. Here, using antibody landscapes of hemagglutination inhibition titers it was shown that infection or vaccination induced a broad increase in titers towards preceding viruses^11,13^. Based on the antigenic distance it was calculated that the back boost extended beyond cross-reactivity^11,13^. However, this mechanism was never validated by antigen binding at a clonal level or shown for any other infectious agents. To our knowledge, we present the first data on a back boost of IgG clones towards a different virus family and substantiate this mechanism with clonal reactivity.

For influenza, back-boost is considered to have a positive contribution to vaccine response by helping maintain immunity towards a broad range of influenza viruses and allowing improved coverage by preemptive vaccination updates^11^. Contrastingly, in OAS, clones initiated by prior infections are shown to negatively impact *de novo* type-specific response and worsen clinical outcome^8–10,13,38^. Whether the SARS-CoV-2-specific response is hampered by back-boost of seasonal clones, similar to what has been described for OAS, is of interest. Immune dominance of IgG reactivity towards seasonal CoV and the late peak SARS-CoV-2 response in these severe COVID-19 patients, 4 weeks after onset of clinical symptoms, suggests the initiation of the SARS-CoV-2 response may be hampered by proliferation of seasonal CoV-specific clones.

In summary, the detailed insights in kinetics and cross-reactivity patterns of N and S-reactive IgG presented in this study will aid in the interpretation of serological assays and expands our understanding of how the humoral immune system responds towards a novel CoV. Also, it shows that studies on in-depth functional characterization of the boosted or newly induced antibody clones after SARS-CoV-2 infection, e.g. VN and ADE assays, are warranted. These data are critical to identify correlates of protection and drivers of immunopathology for COVID-19 and may provide key insights for efficacy and safety testing of future vaccines. Furthermore, the back boost of seasonal CoV IgG shows the immunological background of individuals needs to be considered as an important factor in assessing the quality and quantity of a newly initiated response towards SARS-CoV-2, either by infection or vaccination.

## Data Availability

All data is available for review on request.

## Acknowledgements

This work has received funding from the European Union’s Horizon 2020 research and innovation program under grant agreements No. 874735 (VEO) and No. 101003589 (RECoVER) and the Netherlands Organisation for Health Research and Development (ZONMW) grant agreement 10150062010008. We kindly acknowledge Felicity Chandler BSc for technical assistance, Ron A.M. Fouchier PhD and Georges M.G.M. Verjans PhD for scientific discussions (all Erasmus MC, Rotterdam, the Netherlands) and Berend-Jan Bosch PhD for supplying SARS-CoV-2 and OC43-S_ECTO_ antigens (University of Utrecht, the Netherlands).

