## Supplementary figures for "Severe COVID-19 patients display a back boost of seasonal coronavirus-specific antibodies"

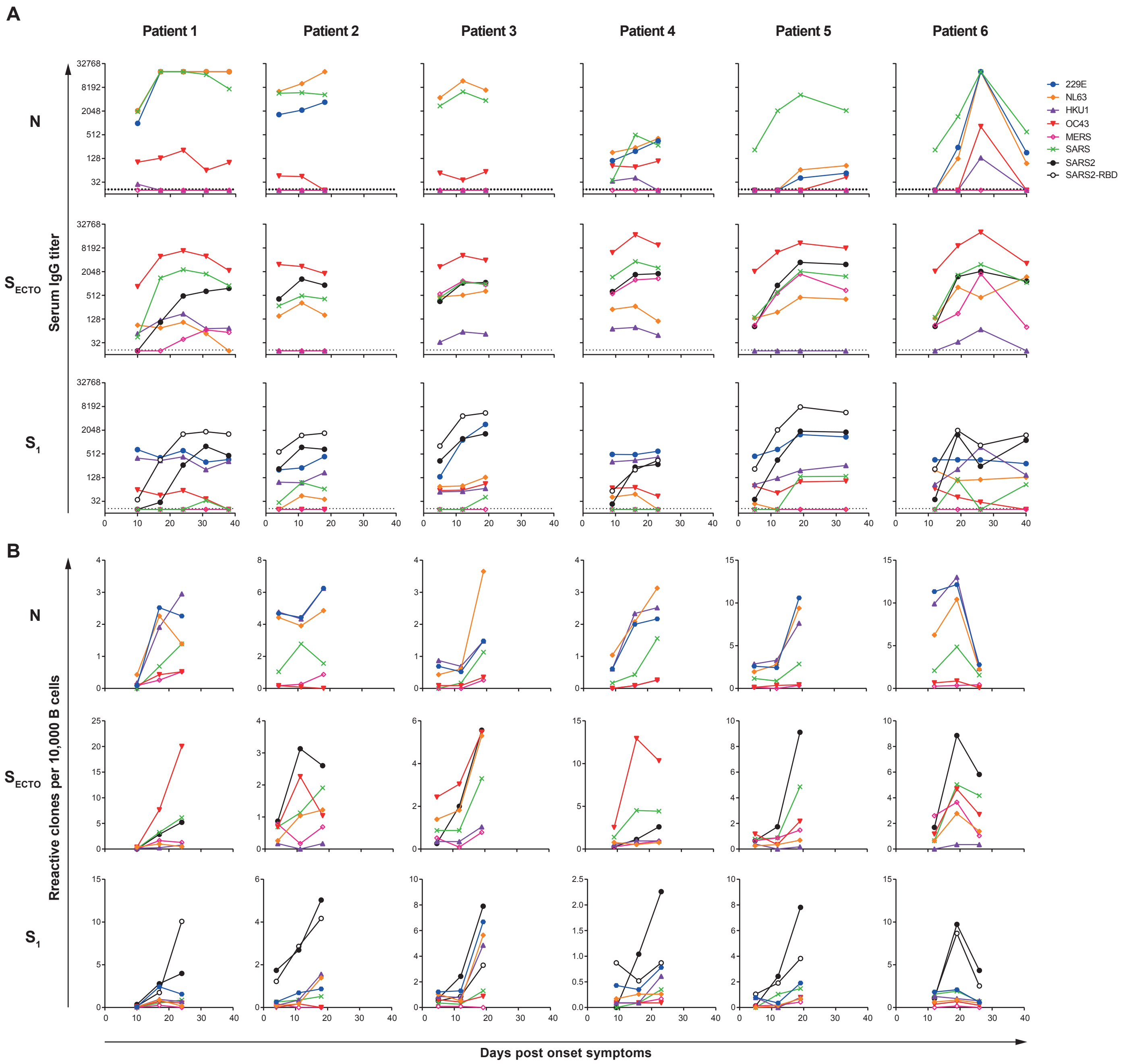


**Supplementary figure 1. Longitudinal paired serum and in vitro stimulated B-cell IgG responses towards coronavirus antigens of six severe COVID-19 patients. A)** Serum IgG titers towards seasonal HCoV-229E, -NL63, -HKU1 and -OC43 and emerging MERS-, SARS- and SARS-CoV-2 nucleocapsid protein (N), ectodomain ($S_{ECTO}$) and head domain of spike ($S_1$) were determined on protein microarray. Dotted line represents the lower limit of detection. **B**) Paired peripheral blood-derived B-cells were isolated and oligoclonaly stimulated *in vitro* at limiting density emperically shown to give a clonal response. The normalized B-cell count of coronavirus IgG reactive culture supernatants on protein microarray are shown.

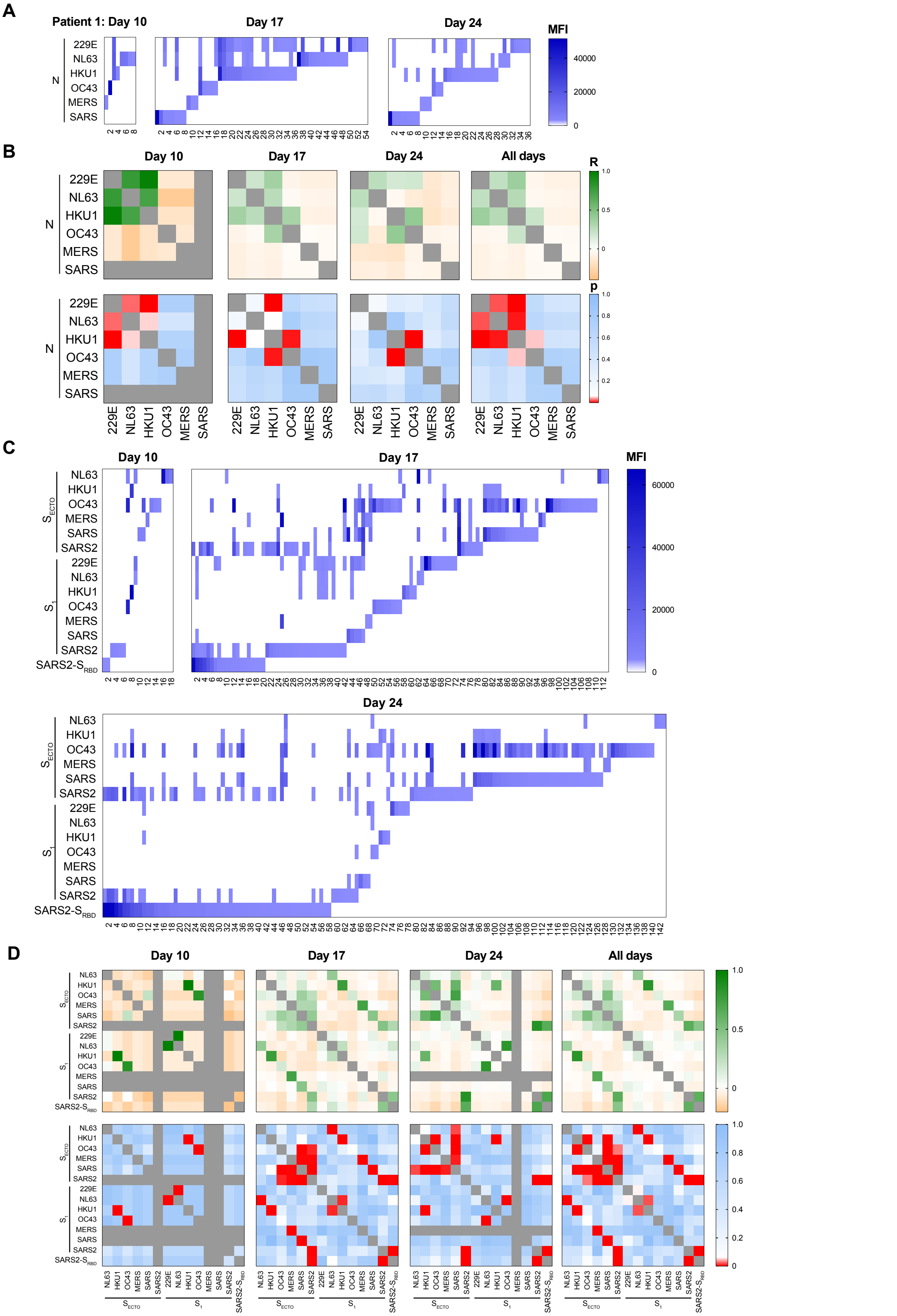


**Supplementary figure 2. Reactivity of IgG expressed by *in vitro* stimulated B-cells at limiting density of patient 1.**

B-cells were isolated from peripheral blood and stimulated in vitro in oligoclonal cultures at limiting dilution to analyze IgG reactivity at the clonal level. A representative patient analyzed at weekly intervals starting at 9 days post onset of symptoms is shown. **A**) Heatmaps of the mean fluorescent intensity (MFI) of clonal IgG reactivity towards N of a panel of coronaviruses. The number of single- and cross-reactive NP-specific B-cells increased over time after SARS-CoV-2 infection. **B**) Heatmaps of the Pearson regression co-efficient (R, top panels) and significance (p, bottom panels) between NP-reactivity of specific B-cell clones shows the level of cross-reactivity at weekly intervals and the clones of all time points combined. Significant positive correlations show a specific clone is likely to cross-react and negative associations shows a clone is likely not to cross-react with the corresponding antigen. **C**) Heatmaps and **D**) regression analysis of $S_{ECTO}$, $S_1$ and $S_{RBD}$ reactive IgG clones from the same patient is shown. Positive correlations are shown in green shades, negative correlations in orange shades. Significant associations are shown in red shades ($p<0.05$).

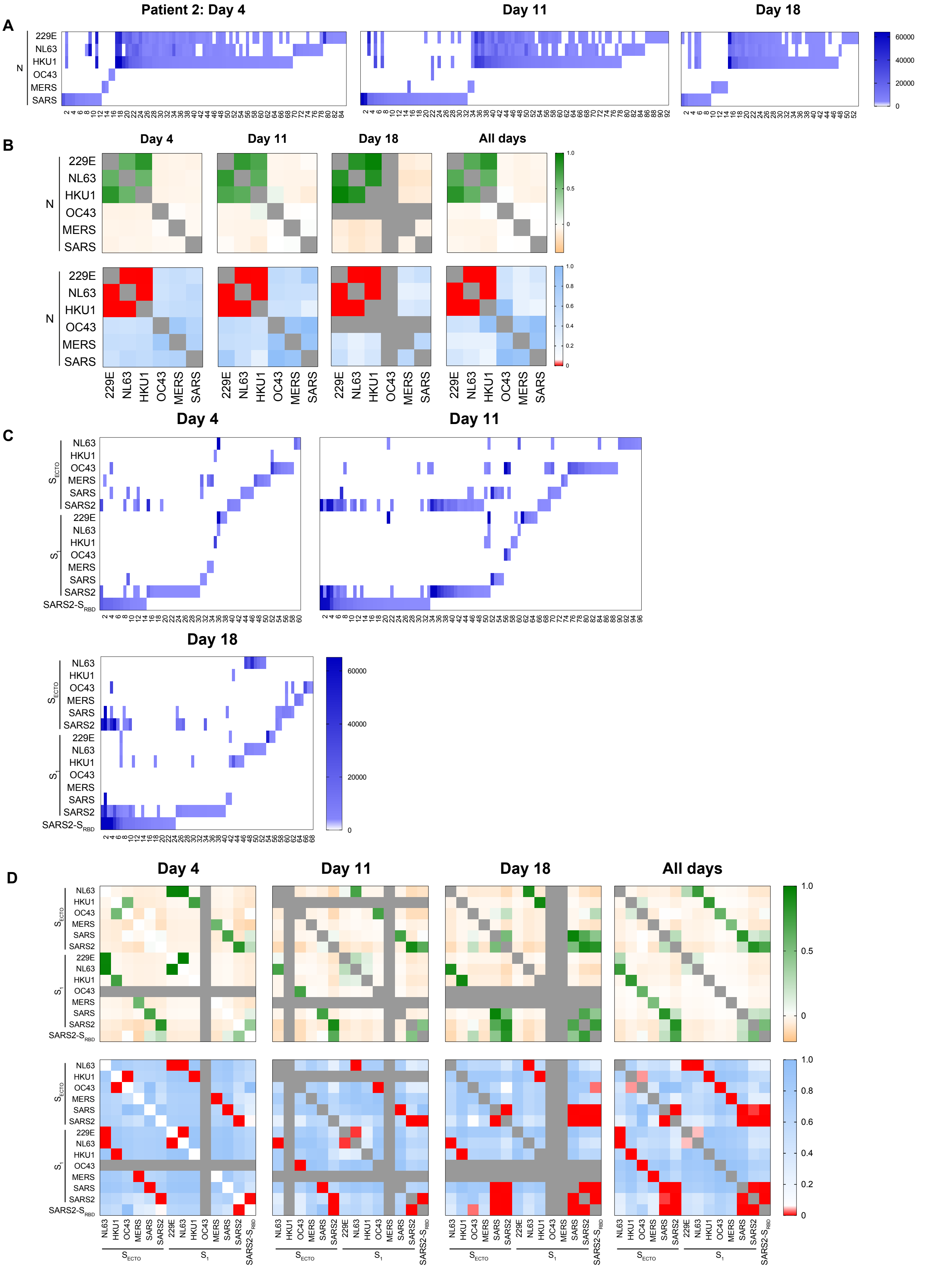


**Supplementary figure 3. Reactivity of IgG expressed by *in vitro* stimulated B-cells at limiting density of patient 2.**

B-cells were isolated from peripheral blood and stimulated in vitro in oligoclonal cultures at limiting dilution to analyze IgG reactivity at the clonal level. A representative patient analyzed at weekly intervals starting at 9 days post onset of symptoms is shown. **A**) Heatmaps of the mean fluorescent intensity (MFI) of clonal IgG reactivity towards N of a panel of coronaviruses. The number of single- and cross-reactive NP-specific B-cells increased over time after SARS-CoV-2 infection. **B**) Heatmaps of the Pearson regression co-efficient (R, top panels) and significance (p, bottom panels) between NP-reactivity of specific B-cell clones shows the level of cross-reactivity at weekly intervals and the clones of all time points combined. Significant positive correlations show a specific clone is likely to cross-react and negative associations shows a clone is likely not to cross-react with the corresponding antigen. **C**) Heatmaps and **D**) regression analysis of $S_{ECTO}$, $S_1$ and $S_{RBD}$ reactive IgG clones from the same patient is shown. Positive correlations are shown in green shades, negative correlations in orange shades. Significant associations are shown in red shades ($p<0.05$).

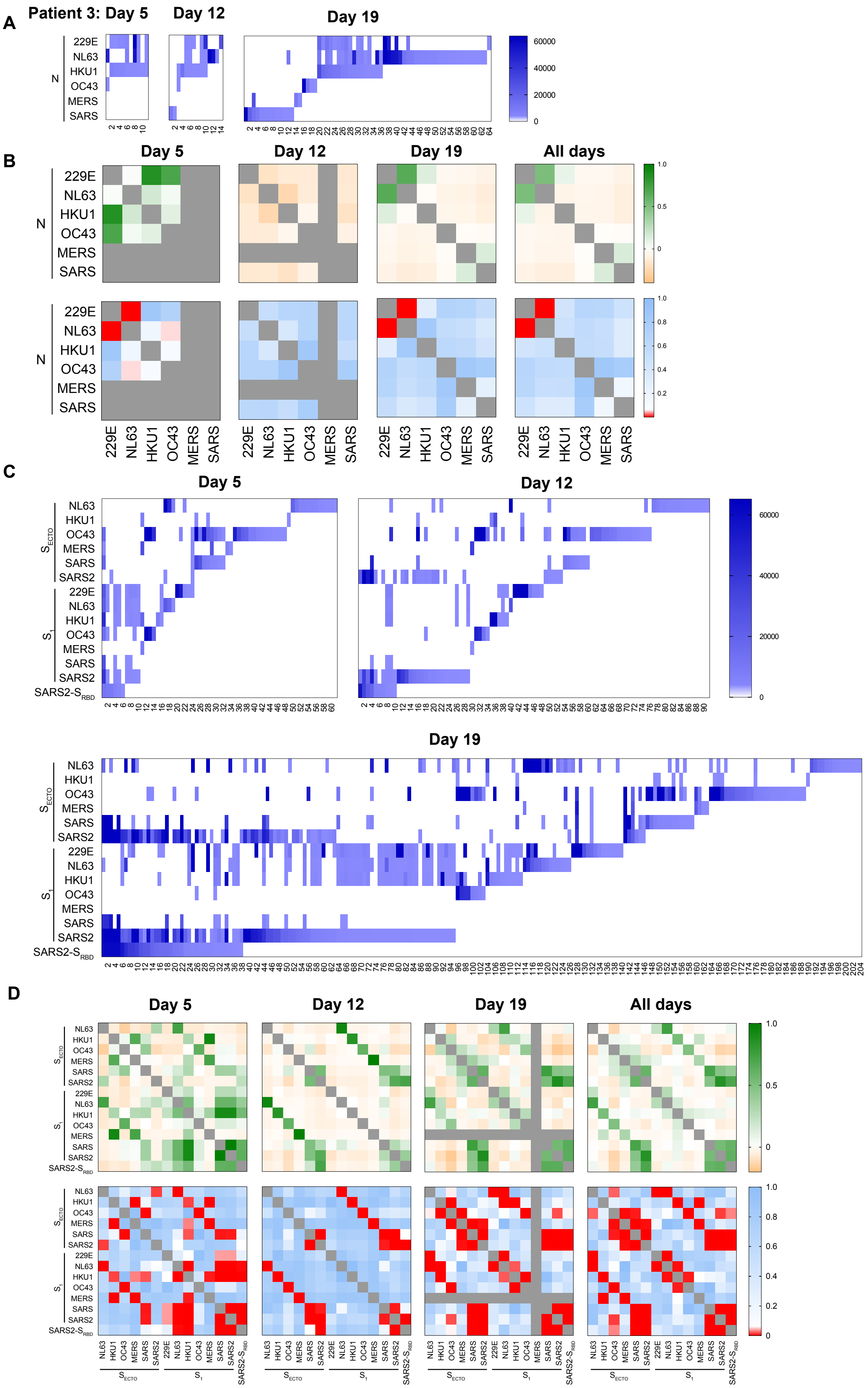


**Supplementary figure 4. Reactivity of IgG expressed by *in vitro* stimulated B-cells at limiting density of patient 3.**
B-cells were isolated from peripheral blood and stimulated in vitro in oligoclonal cultures at limiting dilution to analyze IgG reactivity at the clonal level. A representative patient analyzed at weekly intervals starting at 9 days post onset of symptoms is shown. **A**) Heatmaps of the mean fluorescent intensity (MFI) of clonal IgG reactivity towards N of a panel of coronaviruses. The number of single- and cross-reactive NP-specific B-cells increased over time after SARS-CoV-2 infection. **B**) Heatmaps of the Pearson regression co-efficient (R, top panels) and significance (p, bottom panels) between NP-reactivity of specific B-cell clones shows the level of cross-reactivity at weekly intervals and the clones of all time points combined. Significant positive correlations show a specific clone is likely to cross-react and negative associations shows a clone is likely not to cross-react with the corresponding antigen. **C**) Heatmaps and **D**) regression analysis of $S_{ECTO}$, $S_1$ and $S_{RBD}$ reactive IgG clones from the same patient is shown. Positive correlations are shown in green shades, negative correlations in orange shades. Significant associations are shown in red shades ($p<0.05$).

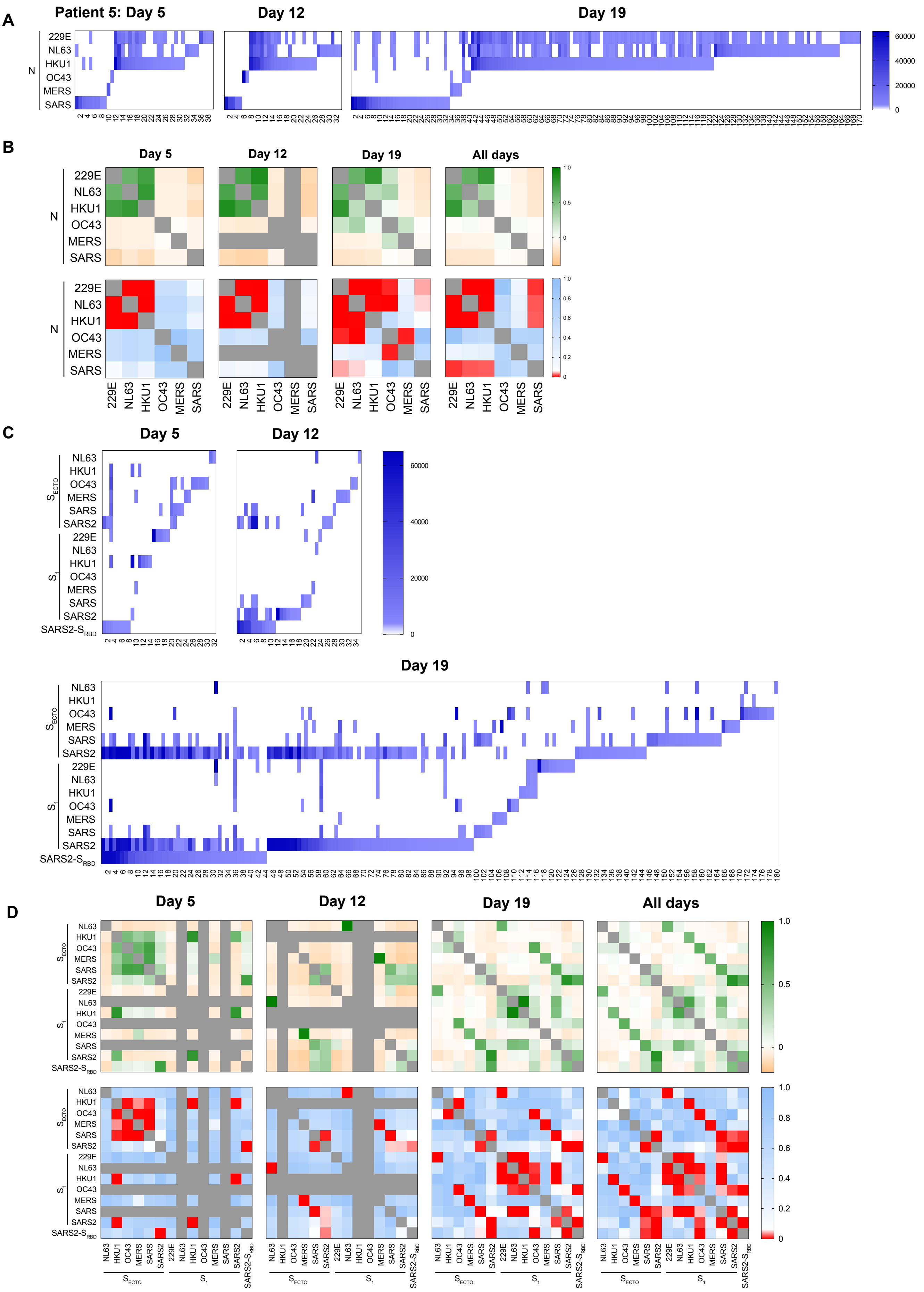


**Supplementary figure 5. Reactivity of IgG expressed by *in vitro* stimulated B-cells at limiting density of patient 5.** B-cells were isolated from peripheral blood and stimulated in vitro in oligoclonal cultures at limiting dilution to analyze IgG reactivity at the clonal level. A representative patient analyzed at weekly intervals starting at 9 days post onset of symptoms is shown. **A**) Heatmaps of the mean fluorescent intensity (MFI) of clonal IgG reactivity towards N of a panel of coronaviruses. The number of single- and cross-reactive NP-specific B-cells increased over time after SARS-CoV-2 infection. **B**) Heatmaps of the Pearson regression co-efficient (R, top panels) and significance (p, bottom panels) between NP-reactivity of specific B-cell clones shows the level of cross-reactivity at weekly intervals and the clones of all time points combined. Significant positive correlations show a specific clone is likely to cross-react and negative associations shows a clone is likely not to cross-react with the corresponding antigen. **C**) Heatmaps and **D**) regression analysis of $S_{ECTO}$, $S_1$ and $S_{RBD}$ reactive IgG clones from the same patient is shown. Positive correlations are shown in green shades, negative correlations in orange shades. Significant associations are shown in red shades ($p<0.05$).

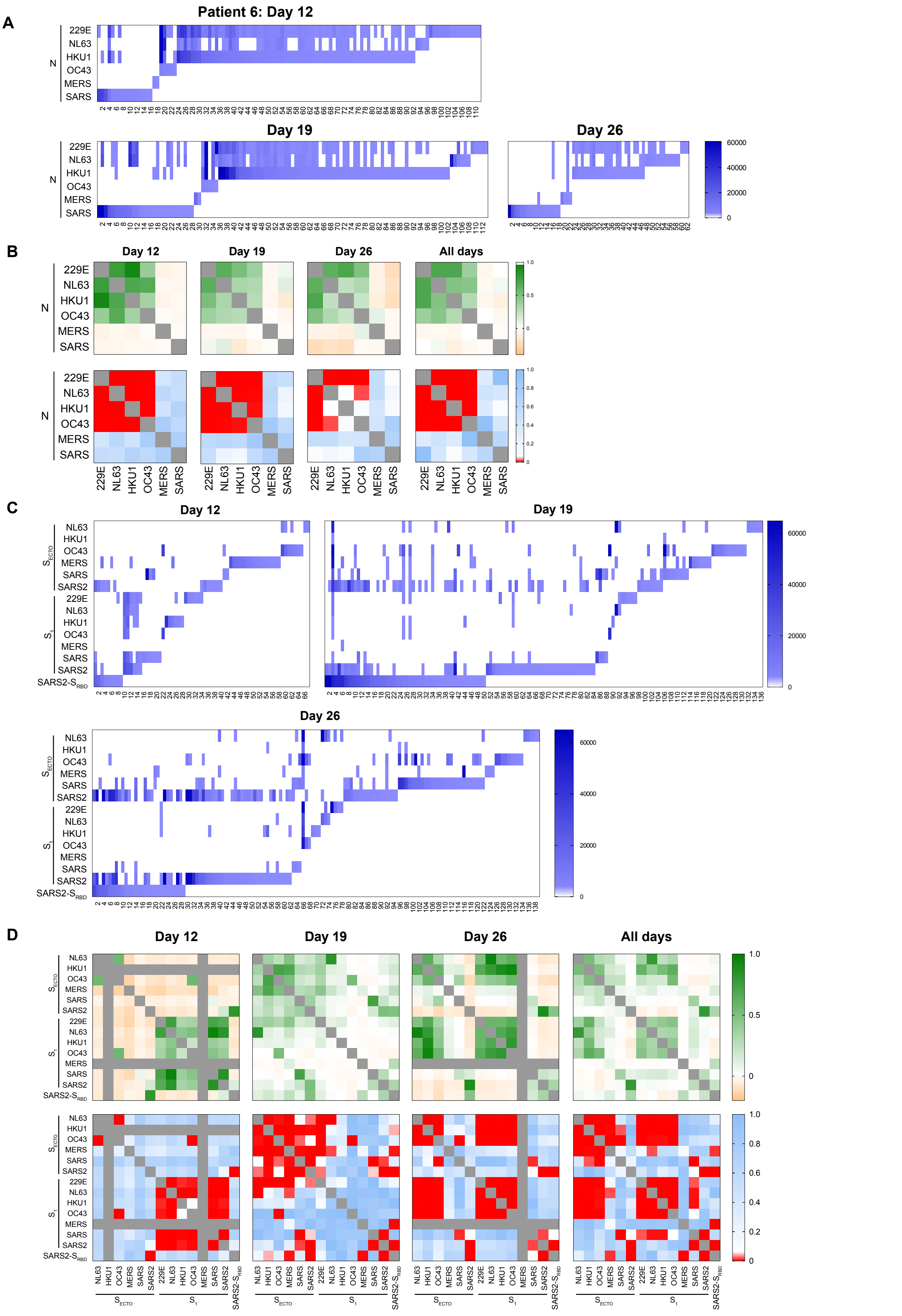


**Supplementary figure 6. Reactivity of IgG expressed by *in vitro* stimulated B-cells at limiting density of patient 6.** B-cells were isolated from peripheral blood and stimulated in vitro in oligoclonal cultures at limiting dilution to analyze IgG reactivity at the clonal level. A representative patient analyzed at weekly intervals starting at 9 days post onset of symptoms is shown. **A**) Heatmaps of the mean fluorescent intensity (MFI) of clonal IgG reactivity towards N of a panel of coronaviruses. The number of single- and cross-reactive NP-specific B-cells increased over time after SARS-CoV-2 infection. **B**) Heatmaps of the Pearson regression co-efficient (R, top panels) and significance (p, bottom panels) between NP-reactivity of specific B-cell clones shows the level of cross-reactivity at weekly intervals and the clones of all time points combined. Significant positive correlations show a specific clone is likely to cross-react and negative associations shows a clone is likely not to cross-react with the corresponding antigen. **C**) Heatmaps and **D**) regression analysis of $S_{ECTO}$, $S_1$ and $S_{RBD}$ reactive IgG clones from the same patient is shown. Positive correlations are shown in green shades, negative correlations in orange shades. Significant associations are shown in red shades ($p<0.05$).
